# Food insecurity during COVID-19: A multi-state research collaborative

**DOI:** 10.1101/2020.12.01.20242024

**Authors:** Meredith T. Niles, Emily H. Belarmino, Farryl Bertmann, Erin Biehl, Francesco Acciai, Anna Josephson, Punam Ohri-Vachaspati, Roni Neff

## Abstract

The COVID-19 pandemic has had profound impacts on the global food system, supply chain, and employment, which, in turn, has created numerous challenges to food access and food security. Early exploratory studies suggest significant increases in food insecurity in the United States. Comprehensive longitudinal research across multiple locations is needed to understand the range of impacts and responses at the household level and to improve preparedness for future events. This protocol paper outlines the formation of the <u>**N**</u>ational <u>**F**</u>ood <u>**A**</u>ccess and <u>**C**</u>OVID research <u>**T**</u>eam (NFACT), a collaborative, interdisciplinary, multi-state research effort that will utilize common measurement tools, codebooks, code, data aggregation tools, and outreach materials to collectively examine and communicate the effect of COVID-19 on household food access and security. NFACT is led by an executive committee of researchers from four institutions, with additional NFACT collaborating institutions across more than a dozen states. A survey was developed by the NFACT executive team in March 2020, with additional refinements in May 2020, using both existing validated questions and new original questions, which were piloted and validated in Vermont. The project provides suggestive guidance for recruitment, and is designed to allow each study site to adopt recruitment strategies that meet their budget and needs. Primary outcomes of interest include food security status, employment status, food access challenges and concerns, dietary intake, and use of food assistance programs. Additional outcomes assess emotional eating, stigma, COVID-19 perceptions and experiences, and pro-environmental purchasing behaviors. This protocol and the establishment of NFACT provide important advancements in COVID-19 and food security research by generating harmonized data and assessing comparable outcomes across geographies and time. The collaborative, open-source approach makes research tools available to teams who might not have the resources to design their own tools, and can enable streamlined data collection, large-scale comparative analyses, and cost savings through reduced administrative tasks. The project has contributed to building new networks between and within states. Enabling facilitation and implementation of instruments in study sites has provided flexibility and meaningful opportunity for local stakeholder engagement and relevant outreach for informed public health decision-making.

## 1. Background

COVID-19 and the social distancing policies implemented to slow its spread have had profound impacts on the global food system and supply chains. Soaring unemployment and underemployment, disproportionately affecting women, low-income communities and people of color (Kocchar 2020 National Public Radio 2020), have created significant food affordability and access challenges. Early evidence from the United States suggests that the food insecurity rate has risen across the country, reaching levels unprecedented in recent history (Niles et al. 2020a, Wolfson and Leung, 2020). The existing research to understand the impact of COVID-19 on food security outcomes is limited, albeit likely to grow. One national survey conducted at the end of March 2020, asking respondents about their food security in the last three months, found adult food insecurity rates as high as the 40% range in some states (Fitzpatrick et al. 2020). However, that work did not explore the factors related to these outcomes, or the sociodemographic characteristics of food insecure respondents. Wolfson and Leung (2020) also surveyed a national sample of low-income individuals in March 2020, finding that 36% of respondents were experiencing food insecurity. Additional reports have documented the increase in food insecurity among households with children, finding that 40% of these households experienced food insecurity in April 2020 (Bauer 2020). Other surveys and polls, such as the U.S. Census Household Pulse Survey (2020), have also shown extraordinary food insecurity rates in the early weeks of the pandemic. A population-level survey in Vermont, conducted between March and April 2020, found that food insecurity had risen 33% since the start of the COVID-19 pandemic (Niles et al. 2020a). Other recent studies have suggested that diet quality during COVID-19 can also change significantly, with some evidence indicating increases in diet quality (Ruiz-Roso et al. 2020, Rodriguez-Perez et al. 2020), and other popular press indicating decreased diet quality (New York Times 2020).

These early efforts to document trends in food insecurity have been critical to support early policy and program responses. At the same time, with the exception of Niles et al. (2020a), these studies generally have focused on food affordability and do not examine multiple dimensions of food security jointly (such as food access, dietary shifts and nutrition, availability and acceptability), nor interrelationships with other COVID-related and contextual factors (such as social distancing behaviors, attitudes, and policy). With projections that the COVID-19 pandemic will continue to affect lives for many more months (if not years), it is critical to understand its effects on food access and security broadly defined, and over time across diverse population groups in order to facilitate well-targeted responses and improve planning for the future.

This project aims to meet this need by establishing a national research collaborative, developing shared survey instruments, and building tools for data aggregation and management. We have convened a collaborative — the National Food Access and COVID research Team (NFACT) — which will use common assessment measures for food access and food security, by (1) implementing the NFACT survey (Niles et al. 2020b,c) across national and subnational (e.g. state, city) samples, and (2) integrating the data with location-specific information on COVID-19 prevalence, social distancing guidance and rules, and relevant policy decisions to comprehensively examine food access and food security outcomes, as well as the sociodemographic factors associated with them.

## 2. Methods/Design

### 2.1 Study Aim

This project aims to meet the need for rigorous documentation of food insecurity during the COVID-19 pandemic by establishing a national research collaborative, developing shared survey instruments to facilitate data collection across population groups, and building tools for harmonized data aggregation and management. The study further aims to document the trends in food access and security over time, nationally and across multiple regions. To monitor these trends at the national level, we will collect data from five repeated cross-sectional national samples, between July 2020 and August 2021. In addition, 16 additional study sites (at the time of publication) will enable understanding of these issues in greater depth across more than a dozen states. Our intent is to provide timely and comprehensive analyses to assess the complex factors associated with food access, food security, and diet outcomes during the COVID-19 pandemic, as well as the potential impact of policies, interventions, and programs designed to blunt the adverse effects of the pandemic on household livelihood and wellbeing (e.g. expansion of food assistance programs, federal stimulus checks, etc.). Additionally, NFACT provides open access to the tools to enable use of similar measures for comparable data, which reduces barriers to research among diverse academic and stakeholder groups interested in gathering household data at local or state levels.

### 2.2. Study Design, Recruitment, and Participant Characteristics

#### 2.2.1 Survey Development

A survey was developed by the research team in March 2020 (Niles et al. 2020b), with additional refinements in May 2020 (Niles et al. 2020c), to measure food access, food security, food purchasing, food assistance program participation, dietary intake, perceptions of COVID-19, and individual social distancing behaviors, as well as household and individual sociodemographics. The survey was explicitly designed to measure key outcomes (e.g. food access, food security, food purchasing, and dietary intake) both prior to the COVID-19 outbreak (dated as March 11th, the day the World Health Organization declared a global pandemic (WHO 2020) and since the pandemic began. The first version was developed in consultation with key stakeholders in the state of Vermont, where it was first implemented, and drew from the existing literature on food security and food access. When possible, validated questions and instruments were used. The survey was piloted in Vermont, with 25 eligible (18 or older) residents in late March, and validation methods (e.g. Cronbach alpha, factor analysis) were used to test the internal validity of questions with key constructs (alpha > 0.70).

Following deployment of the first version of the survey in Vermont, refinements to the original survey tool were made based on the feedback received about question wording and interpretation. New questions were also added related to use of food assistance programs and their functionality during COVID-19, as well as dietary intake, emotional eating, and pro-environmental behaviors (Niles et al. 2020c). All survey materials are currently publicly available (Niles et al. 2020b, Niles et al. 2020c). Table 1 presents outcomes and measures included in version 2.1 of the survey.

**Table 1.** Primary and Secondary Outcomes based on Survey Version 2.1.

| Primary Outcomes | Source/Instrument |
| --- | --- |
| Food security status: Affordability | USDA six-question module |
| Food security status: Food access | Original newly developed questionnaire |
| Food security status: Food availability | Original newly developed questionnaire |
| Food security status: Food acceptability and quality | Original newly developed questionnaire |
| Employment status and changes | Original newly developed questionnaire |
| Dietary intake | NCI screeners, Yaroch et al. 2012 |
| Use of food assistance programs | Original newly developed questionnaire |

| **Secondary Outcomes** | **Source/Instrument** |
| --- | --- |
| Use, acceptability of food assistance programs | Original newly developed questionnaire |
| Emotional Eating | Intuitive Eating Scale-2 (Tylka and Kroon 2013) |
| Stigma of food assistance programs | Based on Pinard et al. (2017) |
| Pro-environmental purchasing behavior | Original newly developed questionnaire |
| COVID-19 perceptions and experiences | Original newly developed questionnaire |
| Food system implications | Original newly developed questionnaire |

#### 2.2.2 Sampling Approaches

This project primarily utilizes two sampling approaches: pre-existing survey panels and opportunity/convenience sampling. NFACT collaborators are encouraged to choose the approach that best meets their research goals and resource constraints.

##### Pre-existing survey panel sampling

The market research approach uses a panel of respondents within a given region (e.g. city, state, country). The survey is administered online. To support rapid survey deployment, we have programmed the survey for the Qualtrics survey platform (Provo, UT), but other survey platforms may be used. We developed two sampling strategies for achieving a representative sample, although teams have the option to adapt these to their own needs.

###### General population sample, representative of the target population with respect to income, race, and ethnicity

This sample is achieved by matching sample recruitment quotas to the income, race (specifically White, Black of African American, American Indian and Alaska Native, Asian, Native Hawaiian or Other Pacific Islander, and Two or more races), and ethnicity (Hispanic, non-Hispanic) population profile in the American Community Survey (ACS). We advise six income categories to capture household income. We also encourage consideration of the sample profile with respect to age and sex. If the Qualtrics platform is used, standard age and sex quotas can be applied.

###### Population sample with heightened risk for food insecurity

This sample is achieved by ensuring recruitment quotas that over represent low-income, minority, or low-education households. As with the general population sample, we encourage consideration of the age and sex distribution of the sample.

##### Opportunity/Convenience sampling

Following the strategy employed in Vermont in March/April 2020 with the V1 survey (Niles et al. 2020a), the opportunity sampling approach employs multiple recruitment methods including the use of local, regional, or statewide listservs; targeted social media campaigns; non-profit, industry, state agency, or other stakeholder listservs; and media publicity. We encourage other recruitment strategies relevant to the target population (individuals over 18 years of age who reside within the geographic area under investigation). The survey can be implemented in any format (e.g. online, phone, or in-person administration).

#### 2.2.3 Study Setting

The survey will be implemented at a minimum at the following locations: Alabama, Arizona, California-Bay Area, Greater Chicago area, Connecticut, Maine, Maryland, Massachusetts, Michigan (Detroit area), New Mexico, New York City, New York State (Albany region), New York State (additional regions), Utah, Vermont, Washington, Wisconsin as well as through national samples.

#### 2.2.4 Ethical Approval and Data Sharing

All study sites obtained Institutional Review Board approval at their own institutions prior to implementing the survey or recruitment of subjects. Data will be shared amongst NFACT collaborators facilitated through the use of data sharing agreements, common codebooks and code (NFACT 2020), and an aggregated NFACT database.

## 3. Primary Outcomes

The primary goal of the study is to understand the changes, if any, to the food insecurity status of respondents since the outbreak of COVID-19, and over time, in the case of repeated measures, as well as the key factors associated with food insecurity outcomes. Unlike most other current assessments of food insecurity during COVID-19, the study assesses multiple dimensions of food insecurity beyond affordability, including access, availability, safety, quality and acceptability. Key factors that can be evaluated in relation to food insecurity include: 1) sociodemographic factors (e.g. age, race, gender, income-level, household composition and size); 2) employment status and recent changes (e.g. job loss or disruption); 3) dietary intake and purchasing changes since COVID-19; and 4) use of food assistance programs and policies since COVID-19. Additional outcomes include the use, acceptability, and functionality of food assistance programs and policies, stigma of food assistance program use, purchasing behavior changes since COVID-19, perceptions of and experiences with COVID-19, emotional eating experiences since COVID-19, and the broader food system impacts of COVID-19, as evidenced through qualitative questions. All primary and secondary outcomes are listed in Table 1.

## 4. Additional Data

The study will also use additional data from outside sources to account for key differences across the states under investigation. These additional data sources include COVID-19 prevalence data, policy/social distancing responses (Johns Hopkins University 2020), and other contextual factors separate from COVID-19.

## 5. Statistical Analysis and Sample Size Calculation

Each study site using the survey will perform its own analysis for purposes of local reporting and insight. They are encouraged to use statistical weighting to match key population-level Census or ACS characteristics. Sample size calculations indicate that for the populations of all the regions of interest, a sample size of greater than 600 in each region would provide a +/-4% margin of error with 95% confidence. Many states and the national survey have opted for sample sizes of 1000, which can provide close to a +/- 3% margin of error with 95% confidence. Code for data quality checks, data cleaning, and variable transformations will be developed for multiple statistical programs and added to a central repository on GitHub (NFACT 2020).

The data from all study sites meeting quality criteria will be aggregated into a common dataset, facilitated by the use of common codebooks and data quality checks based on internal consistency (e.g. household composition and food program participation), and weighting strategies. The data will be analyzed using multiple statistical approaches including ANOVA and chi-square, linear and logistic regressions, hierarchical random effects models to control for repeated measures and geographical correlation (e.g. zipcode, county and/or state), and structural models.

## 6. Project Management and NFACT

This effort establishes NFACT, a collaborative of researchers across the United States committed to timely, collective, and impactful research focused on food access and security during COVID-19. Dr. Meredith Niles of the University of Vermont initiated and serves as overall leader of the project. In addition, NFACT is led by an executive committee, consisting of four founding institutions: The University of Vermont, Johns Hopkins University, Arizona State University, and the University of Arizona. These four institutions and their respective principal investigators (PI) initially designed the survey instrument and provided financial resources and labor for the establishment of the survey instrument, coding, or collaborator support tools (e.g. templates, codebooks). Activities led by the executive committee include database management, data aggregation, decision-making efforts, and communication through bi-monthly calls. As NFACT made their materials open access, presented their materials to national working groups and practice groups, and invited collaboration across the country to facilitate comparative analyses, additional collaborators from 17 study sites have joined NFACT. Each NFACT collaborating institution is led by a PI or multiple PIs, who will independently implement the survey using the above-mentioned strategies, obtain Institutional Review Board approval, govern their data use, lead analyses, and spearhead local communication efforts designed to reach key food security stakeholders.

NFACT collaborators are united in common instruments and tools to aggregate and harmonize data and analysis, providing timely communication of results to key shareholders through commonly-shared brief templates, and describing the impact of COVID-19 on food security and access across diverse regions. Aligned with the project’s open access ethos, NFACT leadership does not place specific requirements on survey users, nor perform oversight (except when data will be aggregated). NFACT does request communication, data sharing, and participation in collaborative discussions. Others with the resources to perform the survey are encouraged to reach out to explore joining the collaborative.

NFACT also acknowledges the disproportionate impact that COVID-19 has had on black, indigenous, and people of color, including both for being more severely affected or dying from COVID-19 (Fortuna et al. 2020, Laster Pirtle 2020, Laurencin and McClinton 2020), as well as food insecurity impacts arising from COVID-19 (Wolfson and Leung 2020). These impacts are the result of how systemic racism directly impacts public health nutrition (Odoms-Young and Bruce 2018). COVID-19 has further amplified structural inequalities especially among black, indigenous, and people of color, who acutely suffer from the shortcomings of our conventional food system (Webb Hooper et al, 2020). Given these factors, NFACT is committed to adopting a racial equity approach (Bread for the World 2019), through which we will build and maintain partnerships with community stakeholders and key policymakers to inform our survey, recruitment, dissemination of results, and policy advocacy in an effort to amplify equity, diversity, and inclusion.

## 7. Discussion

The NFACT collaboration, with its set of common instruments across 17 study sites, aligns with public health research recommendations to utilize common measures (Khan et al. 2009). Given the challenges that the COVID-19 pandemic poses for academic research, there are several important practical and operational elements to note related to this study. First, as initially developed, the survey is implemented online. This was necessary at the beginning of COVID-19 because of social distancing measures, remote and home office work, and for the benefit of timeliness, facilitated through streamlined data collection. At the same time, an online survey format may reduce access for some populations, especially those in rural regions without reliable internet, older adults, and those with very low incomes, who are homeless, or have lower literacy. Language barriers may also impede participation. However, the survey instrument and method do not require online administration, and future surveys can be conducted via telephone or in-person. Adaptation and translation of the survey for populations with limited English proficiency has been pursued by some collaborating institutions.

The survey will be implemented at different times in different study sites, and is currently offered in both English and Spanish. In addition to the explicit aim of gaining diverse snapshots on the pandemic’s effects, further differentials result from the realities of multiple institutional approvals necessary, and different stakeholder engagement processes in various states. We intend to account for these differences in our data aggregation process by noting the date a survey was taken in a given region, and including additional details on the prevalence and policy information on the current conditions of the pandemic in a region on that day. Indeed, even if all survey sites deployed on the same day, the on-the-ground situation would look very different across these places, so the results would still not provide fully parallel information about the relation between COVID-19 and food security.

Regardless of the potential challenges of conducting survey research during COVID-19, there are many benefits of the structure of NFACT and its commitment to collaborative open science. Through the NFACT structure, we have streamlined data collection, reduced redundancy of administrative efforts, provided common instruments and communication tools to facilitate comparative research, and enabled a process and structure to collect data across diverse regions to better understand the challenges of multiple populations. By enabling local facilitation and implementation of instruments in specific regions, PIs and individual institutions can adapt materials to their own community and stakeholder needs, making results relevant and useful for decision-making and policy implementation. Furthermore, the NFACT structure enables access to instruments and materials that may be unavailable due to limited resources at some institutions. The collaboration has contributed to reducing costs, especially in challenging economic conditions, through streamlined instruments, codebooks, code, and outreach materials. Moreover, through a collaborative structure, NFACT is also facilitating many new research collaborations and additional projects, as well as building the professional networks of those engaged, which likely will have long term benefits for scientific research.

## Data Availability

The code associated with this article is cited within the manuscript and is available on GitHub.

https://github.com/aljosephson/nfact

## Conflict of Interest

The authors declare that the research was conducted in the absence of any commercial or financial relationships that could be construed as a potential conflict of interest.

## Author Contributions

MTN, EHB, FB wrote the original manuscript. FA, AJ, and MTN provided data curation, code and codebooks for current NFACT operations. All authors provided conceptualization, data curation, funding acquisition, methods, resources, read, edited and approved the final manuscript.

## Availability of data and materials

Code is available on GitHub (NFACT 2020). Publications and briefs will be made available at www.nfactresearch.org.

## Funding

Funding for the establishment of NFACT was made possible through grants to the University of Vermont: College of Agriculture and Life Sciences/Office of the Vice President of Research, Gund Institute for the Environment, and Center for Food Systems Research; Johns Hopkins University: Directed Research Grant Center for a Livable Future; University of Arizona: College of Agriculture and Life Science; Arizona State University: College of Health Solutions.

## Acknowledgements

We would like to acknowledge all of the NFACT collaborating institutions and collaborators, at the time of publication including: Deanne Allegro (Auburn University at Montgomery), Emily Barbour (University of Vermont), Alyssa Beavers (Wayne State University), Young Cho (University of Wisconsin Milwaukee), Lauren Clay (D’Youville College), Anne Dressel (University of Wisconsin Milwaukee), Beth Feingold (SUNY-Albany), Katie Fiorella (Cornell University), Amy Harley (University of Wisconsin Milwaukee), Kaitlyn Harper (Johns Hopkins University), Linnea Laestadius (University of Wisconsin Milwaukee), Katie Martin (Foodshare), John Mazzeo (DePaul University), Jennifer Otten (University of Washington), Giselle Pignotti (California State University San Jose), Stephanie Rogus (New Mexico State University), Joelle Robinson (Johns Hopkins University), Mateja Savoie Roskos (Utah State University), Rachel Schattman (University of Maine), Brinda Sivaramakrishnan (Tacoma Community College), Rachel Weil (The Greater Boston Food Bank), Kathryn Yerxa (University of Maine), Rachel Zack (The Greater Boston Food Bank)

## Notes

### Competing Interest Statement

The authors have declared no competing interest.

### Author Declarations

Each individual institution seeks their own IRB approval through this collaboration. individual IRBs will be detailed in manuscripts.

